# Adverse childhood experiences and lower urinary tract symptoms in adolescence: the mediating effect of inflammation

**DOI:** 10.1101/2024.05.14.24307366

**Authors:** Kimberley Burrows, Jon Heron, Gemma Hammerton, Ana L. Goncalves Soares, Carol Joinson

## Abstract

**Background:** There is evidence that adverse childhood experiences (ACEs) are associated with lower urinary tract symptoms (LUTS) in adulthood, but few studies have explored these associations in adolescence. Little is known about the biological mechanisms that could explain these associations. We examine whether inflammatory biomarkers mediate the relationship between ACEs and LUTS in adolescence.

**Methods:** We used data from 4,745 participants from the Avon Longitudinal Study of Parents and Children on ACEs (10 ‘classical’ ACEs assessed from birth to age 8), LUTS at age 14 (any urinary incontinence (UI), daytime and bedwetting, urgency, nocturia, frequent urination, voiding postponement, and low voiding volume) and inflammatory biomarkers interleukin-6 (IL-6) and C-reactive protein (CRP) measured at age 9. We first examined associations between the (i) ACE score (summed score [scale of 0 to 10] of total ACEs) and LUTS and (ii) inflammation and LUTS using multivariable logistic regression. We evaluated the mediating effects of IL-6 and CRP on the association between the ACE score and LUTS using the parametric g-formula whilst adjusting for baseline and intermediate confounders.

**Findings:** Higher ACE scores were associated with increased odds of LUTS, e.g. a one-unit increase in ACE score was associated with an increase in the odds of any UI (odds ratio [OR] 1·16, 95% confidence interval [CI] 1·03-1·30). Higher levels of IL-6 were associated with increased odds of LUTS, e.g. any UI (OR 1·24, 95%CI 1·05-1·47). There was weak evidence that the associations between ACE score and any UI, daytime wetting, bedwetting, urgency, and frequency were mediated by IL-6 (e.g. any UI OR_natural indirect effect_ 1·03, 95%CI 1·00-1·06, proportion mediated 21%). There was no evidence that CRP was associated with LUTS, nor mediated the association between ACE score and LUTS.

**Interpretation:** This study reports novel findings that could shed light on the biological mechanisms that underlie the link between ACEs and LUTS. Early intervention is needed in childhood to prevent LUTS persisting into adolescence.

**Funding:** Medical Research Council (grant ref: MR/V033581/1: Mental Health and Incontinence).

**Research in context:** *Evidence before this study:* There is growing evidence that adverse childhood experiences (ACEs) are associated with an increased risk of lower urinary tract symptoms (LUTS), but the mechanisms are unknown. One plausible biological mechanism is through ACEs leading to increased inflammation, which has been implicated as a contributing factor for LUTS. We searched PubMed and Google Scholar from March 2023 to January 2024 for studies published in English describing associations between ACEs (search terms: “adverse childhood experiences” OR “ACEs” OR “adversity” OR “adversities” OR “adverse experiences” OR “stressful life events”) and LUTS (search terms: “lower urinary tract symptoms” OR “incontinence” OR “overactive bladder” OR “enuresis” OR “bedwetting” OR “daytime wetting” OR “bladder symptoms” OR “urinary symptoms” OR “urgency”), ACEs and inflammation (search terms: “inflammation” OR “interleukin 6” OR “c reactive protein”), and inflammation and LUTS in populations of children, adolescents and adults (search terms: “child OR childhood”, “adolescent OR adolescence”, “adult”). We did not identify previous cohort studies that have explored the association between ACEs and LUTS during adolescence. Previous studies found that ACEs were associated with LUTS, but they focussed on relatively small samples of adult women, they relied on retrospective recall of ACEs, and one study lacked a control group without LUTS. No cohort studies have to our knowledge examined associations between inflammation and LUTS in adolescence.

*Added value of this study:* To our knowledge, this is the first cohort study to report that exposure to more ACEs between birth and 8 years is associated with an increased risk of subsequent LUTS in adolescence (age 14). We also found that inflammation increases the risk of subsequent LUTS. Finally, we show that the associations between ACEs and LUTS are partially mediated by the inflammation biomarker IL-6.

*Implication of all the available evidence:* Our findings should raise awareness amongst clinicians of the importance of screening for ACEs in children presenting with LUTS. Evidence of biological mechanisms (such as inflammation) linking ACEs to LUTS could lead to the identification of novel translational targets for intervention and potential therapeutic advances in the treatment of LUTS.

## Introduction

Urinary incontinence (UI) is common in childhood, with bedwetting and daytime wetting affecting around 15% and 8% of seven-year-olds, respectively.^1,2^ Although many cases of UI resolve during childhood, a significant proportion persist into adolescence, and new-onset cases also occur.^3^ It is estimated that 2·5% of 14-year-olds experience bedwetting and 2·9% suffer from daytime wetting; other lower urinary tract symptoms (LUTS) such as urgency and nocturia are also common at this age (4·8% and 9·2%, respectively).^3^ Most cases of LUTS in children and young people are functional (non-organic) and can have an adverse effect on wellbeing and mental health.^4,5^

There is growing evidence that psychosocial factors in childhood, including stressful life events and adverse childhood experiences (ACEs) are associated with an increased risk of LUTS^6,7^. ACEs are traumatic experiences including abuse, neglect, and parental substance abuse. Although there is some evidence that certain types of adversity (e.g. sexual abuse^8^) are associated with an increased risk of LUTS, ACEs are often interrelated, making it difficult to isolate the effect of a specific adverse experience^9^. Instead, the total burden of exposure to ACEs during childhood is likely to be more important in determining the risk for subsequent LUTS. Previous research on ACEs and LUTS has mostly focussed on adult women: a cohort study found that ACEs were associated with LUTS;^10^ a case-control study found an association between ACEs and overactive bladder,^11^ and a study based on a convenience sample who presented to a urology clinic found that a greater number of ACEs were associated with a greater number of LUTS.^6^ These studies are however, limited by retrospective recall of ACEs; small sample size,^6,11^ and the lack of a control group without LUTS.^6^ To our knowledge no prospective cohort studies have examined if ACEs are associated with an increased risk of LUTS in adolescence. The potential mechanisms through which exposure to ACEs increases risk of LUTS are also under-explored. One plausible biological mechanism is through ACEs leading to increased inflammation.^12^ Inflammation has also been implicated as a contributing factor for LUTS,^13^ but no cohort studies have examined this in paediatric samples.

In the current study, based on data from the Avon Longitudinal Study of Parents and Children (ALSPAC), we examine prospective relationships between (i) ACEs (from birth to 8 years) and self-reported LUTS at age 14 and (ii) two inflammatory biomarkers (interleukin-6 and c-reactive protein) at age 9 and LUTS. We also examine evidence for a mediating effect of inflammation on the relationship between ACEs and LUTS.

## Methods

We used the Strengthening the Reporting of Observational Studies in Epidemiology (STROBE) cohort reporting guidelines (see STROBE checklist in the Supplementary text).

### Sample

ALSPAC is a prospective, population-based birth cohort study that recruited pregnant women resident in Avon, UK with expected dates of delivery between 1st April 1991 and 31st December 1992. The initial number of pregnancies enrolled was 14,541 resulting in 14,062 live births and 13,988 children who were alive at 1 year of age. When the oldest children were approximately 7 years of age, an attempt was made to bolster the initial sample with eligible cases who had failed to join the study originally resulting in an additional 913 children being enrolled. The total sample size for analyses using any data collected after the age of seven is therefore 15,447 pregnancies, resulting in 15,658 foetuses. Of these 14,901 children were alive at 1 year of age. The phases of enrolment are described in more detail in the cohort profile paper and its update.^14,15^ The study website (www.bristol.ac.uk/alspac) contains details of all the data that is available through a fully searchable data dictionary and variable search tool: http://www.bristol.ac.uk/alspac/researchers/our-data/. Ethical approval for the study was obtained from the ALSPAC Ethics and Law Committee and the Local Research Ethics Committees. Informed consent for the use of data collected via questionnaires and clinics was obtained from participants following the recommendations of the ALSPAC Ethics and Law Committee at the time. Consent for biological samples has been collected in accordance with the Human Tissue Act (2004).

The eligible sample included children who attended the “Focus@9” clinic at the mean age of 9·9 years (SD 0·32; hereafter referred to as age 9 years) and provided a blood sample which was assayed for IL-6 and CRP (N = 4,745). Complete data for inflammatory biomarkers, ACEs, LUTS and confounders was available for 1,399 children. The participant flowchart is presented in Figure 1.

**Figure 1.**
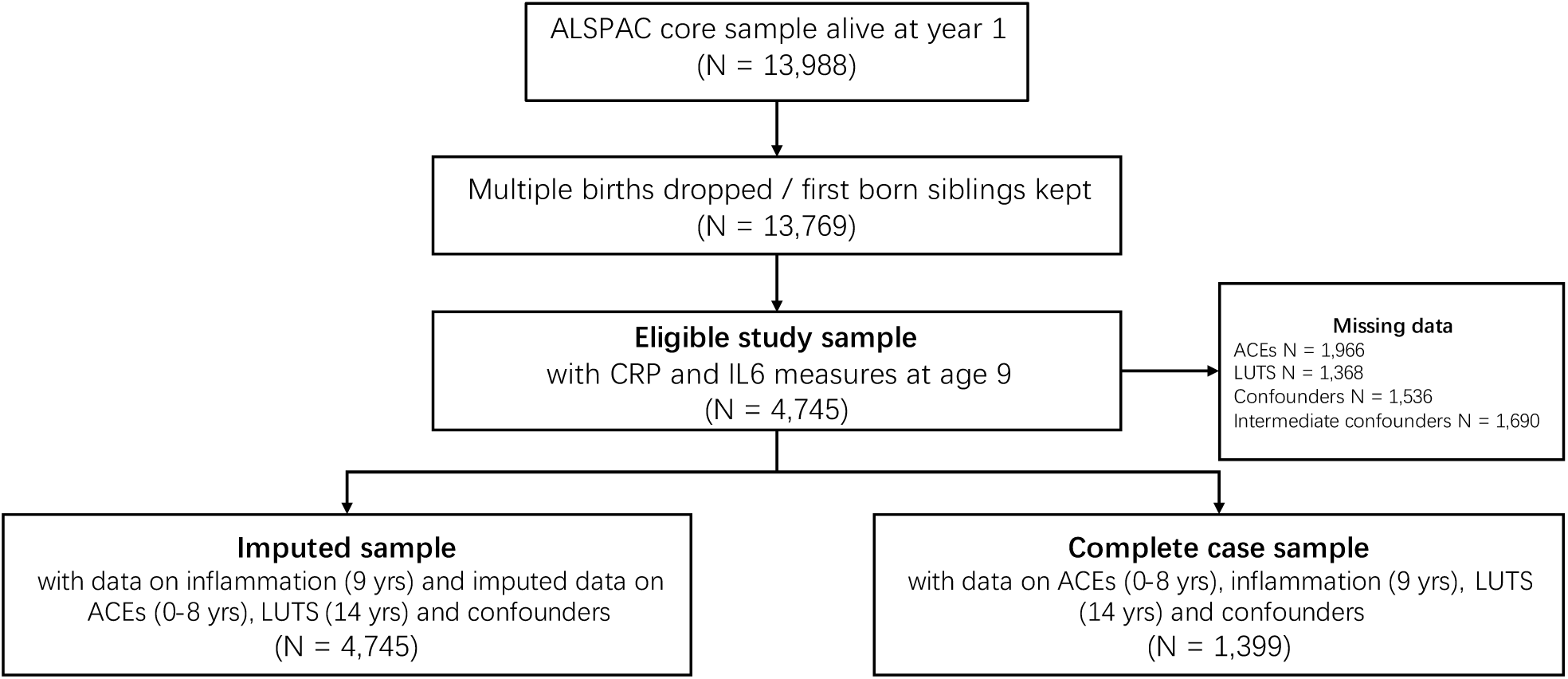
Flow chart of inclusion and exclusion of ALSPAC participants into the study sample ALSPAC Avon Longitudinal Study of Parents and Children; CRP C-reactive protein; IL-6 Interleukin-6; ACEs adverse childhood experiences; LUTS lower urinary tract symptoms.

### Outcomes: LUTS

LUTS were assessed at age 14 (mean (SD) 13·9 (0·14) years) via self-report postal questionnaire as previously described.^3^ Briefly, the questionnaire asked the young person about the presence and frequency of LUTS within the past two weeks including daytime wetting, bedwetting, urgency, frequent urination, low voiding volume, voiding postponement, and nocturia. We additionally derived a variable indicating the presence/absence of any UI (either daytime and/or bedwetting). The specific LUTS questions and variable coding are in Supplementary Table 1.

### Exposure: ACEs

Mothers, partners and children were asked 343 questions over 28 time points about the child’s exposure (experienced vs. not experienced) to 10 ACEs up to age 8 years. The ACE score represents the overall burden of ACEs and is the summed score (scale of 0 to 10) of total experiences of the 10 “classical” ACEs: child sexual, physical, or emotional abuse; emotional neglect; substance abuse by the parents; parental mental illness or suicide attempt; violence between the parents; parental separation; bullying, and parental criminal conviction.^16^ ACEs were considered present if the participant had responded to >50% of the questions relating to the specific ACE (<50% led to the participant being coded as missing), and if criteria for the ACE was met at least once by the time the child was 8 years old. Further details of the definition of each ACE are given in Supplementary Table 2.

### Inflammatory biomarkers: IL-6 and CRP

We explored the mediating effects of two biomarkers of inflammation: IL-6 and CRP. These are well-established inflammatory biomarkers. IL-6 is a cytokine that has pro-inflammatory effects, and CRP is an acute-phase protein produced by the liver in response to IL-6 and TNF-α. IL-6 and CRP were measured using clinical chemistry on non-fasted blood samples taken when the study children were aged 9 years. Blood samples were immediately spun and frozen at -80°C. The biomarkers were assayed after a median of 7·5 years in storage with no previous freeze-thaw cycles. IL-6 (pg/mL) was measured by enzyme-linked immunosorbent assay (R&D Systems) and high-sensitivity CRP (mg/L) was measured by automated particle-enhanced immunoturbidimetric assay (Roche). Inter-assay coefficients of variation for both outcomes were <5%.

### Baseline and intermediate confounders

We adjusted for baseline confounders (measured antenatally or at birth) of each of the mediating pathways (exposure-outcome, exposure-mediator, and mediator-outcome) including indicators of socioeconomic status (SES) (maternal education, housing tenure, housing crowding, and marital status), maternal smoking during pregnancy, maternal age at birth of child, parity, ethnicity, birthweight, gestational age, and sex assigned at birth. We also adjusted for child developmental delay (measured at 18 months).

We adjusted for intermediate confounders at age 8 years that may be caused by ACEs and contribute to inflammation and LUTS (in addition to confounding the mediator-outcome associations). These included body mass index (BMI), constipation, and emotional and behavioural problems. We also adjusted for age at blood collection for the inflammatory biomarkers.

Details on the measurement and coding of the confounders is given in Supplementary Table 3.

### Statistical analysis

We used univariable and multivariable logistic regression to examine the relationships between (i) ACE score and LUTS, and (ii) inflammatory biomarkers and LUTS. We used univariable and multivariable linear regression to confirm previous ALSPAC results for ACE score and the inflammatory biomarkers. IL-6 and CRP measures were natural logarithmically transformed due to their skewed distribution.

We used the *gformula* (parametric g-computation formula) command in Stata to examine the mediating effect of the inflammatory biomarkers on the association between ACE score and LUTS (see Figure 2) (see Supplementary text for further information). G-formula is based on the counterfactual framework and allows the decomposition of effects into the total causal effect (TCE), the natural indirect effect (NIE) (via the mediators), and the natural direct effect (NDE) (not via the mediators). G-formula can also estimate marginal odds ratios for mediation effects with a binary outcome whilst incorporating intermediate confounders (see Supplementary text for further information). We used g-formula to decompose the indirect effects through IL-6 and CRP using two models (see Supplementary Figure 1); (i) we specified IL-6 as the mediator whilst omitting CRP because we hypothesised that CRP is on the causal pathway from IL-6 to LUTS (model 1); and (ii) we specified CRP as the mediator and IL-6 as an intermediate confounder preserving the biological pathway between the two biomarkers (model 2). We used a Monte Carlo sample size of 100,000 and we estimated normal-based 95% confidence intervals (CI) using standard errors from 500 bootstrap resamples. We did not hypothesise an interaction between the exposure and mediator and therefore did not include an interaction term within the model. The proportion mediated (PM) was calculated as [OR_NDE_ (OR_NIE_ − 1)] / [OR_NDE_ × OR_NIE_ − 1] × 100, however, this is only interpretable when the TCE, NIE, and NDE have the same direction of effect.

**Figure 2.**
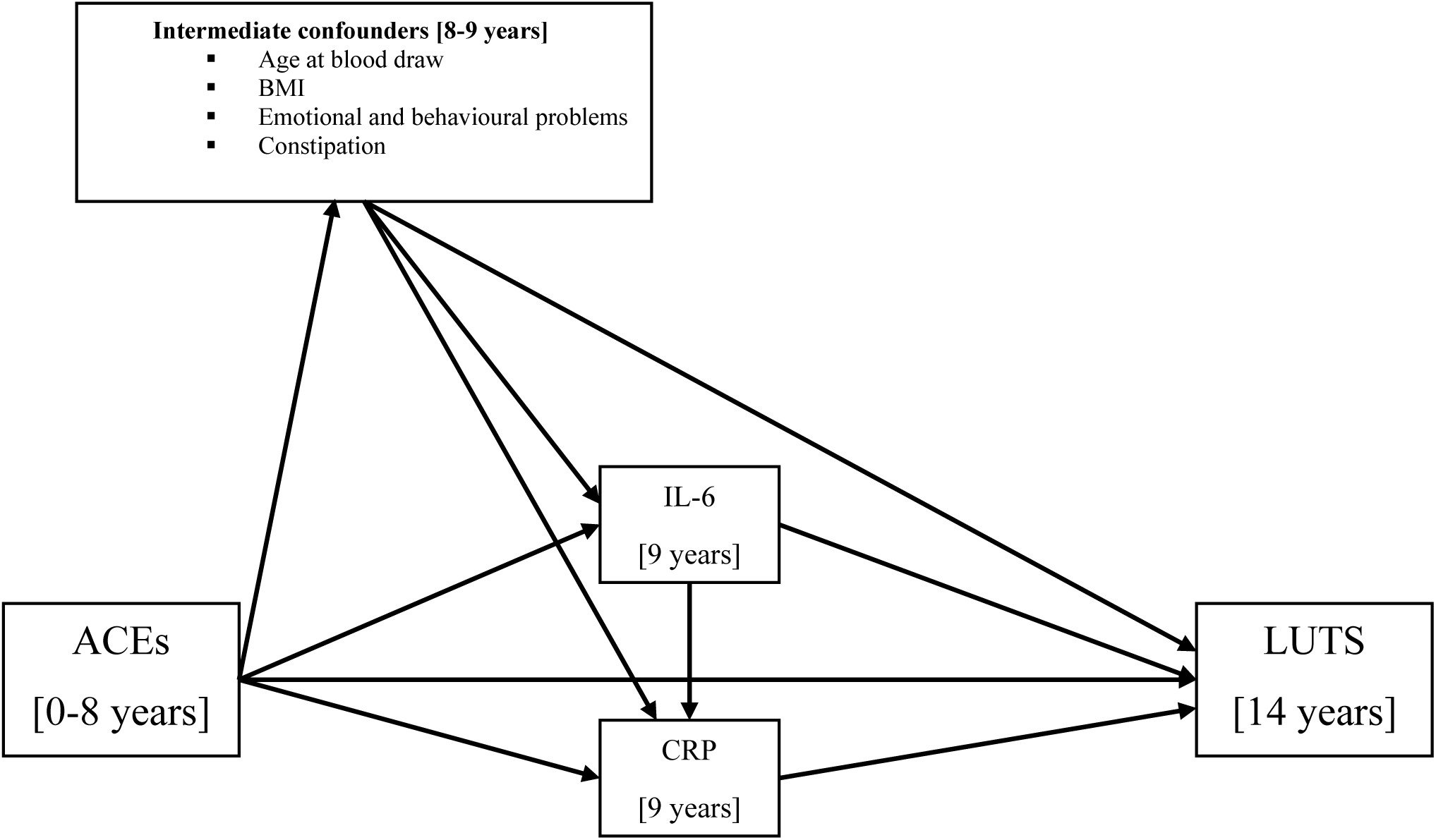
Directed Acyclic Graph (DAG) illustrating hypothesised causal relationships between the exposures, mediators and outcomes Directed acyclic graph (DAG) of the mediation model. Each intermediate confounding path (age at blood draw, child BMI, emotional and behavioural problems, and constipation) was specified separately but are grouped together for clarity. For simplicity baseline confounders are omitted from the DAG as all confounders are proposed to cause all other model variables. ACEs adverse childhood experiences; LUTS lower urinary tract symptoms; BMI body mass index; CRP C-reactive protein; IL-6 Interleukin-6.

Analyses were conducted using Stata version 17 and R version 4·3·0.

### Missing data

We used multivariate imputation by chained equations under the missing at random assumption to impute missing data^17^ in the individual ACEs, LUTS, and confounders up to the sample with complete data on IL-6 and CRP (N = 4,745). We also included auxiliary variables that are considered predictive of the missing values in ACEs and LUTS including SES indicators, adversities reported in the antenatal period and late adolescence, and LUTS variables at age 7 and 9 years (see Supplementary Table 4). There was a lack of good auxiliary variables to enable imputation of missing IL-6 and CRP levels. All binary variables were imputed using logistic regression, categorical (unordered) variables were imputed using polytomous regression, and all continuous variables were imputed using predictive mean matching. ACE score was passively imputed. Missing data were imputed separately in males and females before appending the two datasets together. For both males and females 80 imputed datasets with 50 iterations were created using the mice package (version 3·16·0) in R. Estimates were then combined using Rubin’s rules (see Supplementary text).

## Results

Supplementary Tables 5, 6 and 7 shows the descriptive statistics of study variables for the imputed and complete case samples. Any UI was reported by 5.3% of adolescents, with 4.6% and 4.5% of adolescents experiencing daytime wetting and bedwetting respectively. The most common LUTS were voiding postponement (15.6%) and nocturia (10.5%), while high frequency of urination was the least common (4.3%). Further details of the study sample and variables are given in the Supplementary text.

### Associations between ACEs and inflammation

There was evidence that a higher ACE score was associated with a higher level of IL-6 (expressed as the percentage change in IL-6 per unit increase in ACE score) (1·03 (3%), 95% CI 1·01-1·05 (adjusted); Supplementary Table 8) but not CRP. There was strong evidence of an association between IL-6 and CRP (1·78 (78%), 95% CI 1·72-1·84 (adjusted); Supplementary Table 8).

### Associations between ACEs and LUTS

Higher ACE scores were associated with increased odds of LUTS, e.g. a one unit increase in ACE score was associated with an increase in the odds of any UI (OR 1·16, 95% CI 1·03-1·30 (adjusted)). There were similar associations with all other LUTS, however, the 95% CI’s for daytime wetting and bedwetting cross the null (Table 1). The direction of association for each association was concordant in the complete case analysis (see Supplementary Table 9).

**Table 1.** Associations between adverse childhood experiences score and lower urinary tract symptoms in the imputed data sample (N = 4,745) OR odds ratio; CI confidence interval; UI urinary incontinence; multivariable models adjusted for baseline confounders (sex, birthweight, gestational age, ethnicity, child developmental delay, maternal age, smoking during pregnancy, parity, maternal education, marital status, house tenure, and crowding). Analysis performed in 80 imputed datasets.

| ACE score<br>Outcome | Univariable |  |  | Multivariable |  |  |
| --- | --- | --- | --- | --- | --- | --- |
|  | OR | 95% CI | p-value | OR | 95% CI | p-value |
| UI (any) | 1.20 | 1.08, 1.34 | <0.001 | 1.16 | 1.03, 1.30 | 0.015 |
| Daytime wetting | 1.18 | 1.03, 1.35 | 0.020 | 1.15 | 0.98, 1.34 | 0.081 |
| Bedwetting | 1.21 | 1.05, 1.40 | 0.010 | 1.17 | 0.99, 1.37 | 0.063 |
| Urgency | 1.23 | 1.11, 1.38 | <0.001 | 1.19 | 1.06, 1.34 | 0.004 |
| Nocturia | 1.21 | 1.12, 1.32 | <0.001 | 1.15 | 1.05, 1.26 | 0.004 |
| Frequency | 1.33 | 1.19, 1.50 | <0.001 | 1.29 | 1.12, 1.48 | <0.001 |
| Voiding postponement | 1.13 | 1.05, 1.22 | <0.001 | 1.10 | 1.02, 1.19 | 0.019 |
| Voiding volume | 1.19 | 1.06, 1.34 | 0.003 | 1.15 | 1.00, 1.31 | 0.045 |

### Associations between inflammation and LUTS

A one-unit increase in log IL-6 was associated with an increase in the odds of any UI, daytime wetting, and nocturia in the adjusted models (Table 2). There was weak evidence that IL-6 was associated with reduced odds of voiding postponement (OR 0·89, 95% CI 0·79-1·00). The 95% CIs for associations of IL-6 with the other LUTS crossed the null. The associations were similar in the complete case analysis (see Supplementary Table 10). All 95% CIs crossed the null in the analysis of the association between CRP and LUTS.

**Table 2.** Associations between inflammatory biomarkers and lower urinary tract symptoms in the imputed data sample (N = 4,745) OR odds ratio; CI confidence interval; CRP C-reactive protein; IL-6 Interleukin-6; UI urinary incontinence; multivariable models adjusted for baseline confounders (sex, birthweight, gestational age, ethnicity, child developmental delay, maternal age, smoking during pregnancy, parity, maternal education, marital status, house tenure, and crowding), intermediate confounders (age at blood draw, child BMI, emotional and behavioural problems, and constipation), and ACE score. Analysis performed in 80 imputed datasets; Log is the natural logarithm.

| Inflammation<br>Outcome | Univariable |  |  | Multivariable |  |  |
| --- | --- | --- | --- | --- | --- | --- |
|  | OR | 95% CI | p-value | OR | 95% CI | p-value |
| **Exposure: logIL-6 (pg/ml)** |  |  |  |  |  |  |
| UI (any) | 1.29 | 1.11, 1.51 | 0.001 | 1.24 | 1.05, 1.47 | 0.011 |
| Daytime wetting | 1.50 | 1.21, 1.86 | <0.001 | 1.41 | 1.11, 1.79 | 0.005 |
| Bedwetting | 1.14 | 0.92, 1.42 | 0.232 | 1.11 | 0.89, 1.39 | 0.336 |
| Urgency | 1.18 | 0.99, 1.40 | 0.058 | 1.10 | 0.92, 1.32 | 0.311 |
| Nocturia | 1.24 | 1.09, 1.42 | 0.001 | 1.16 | 1.01, 1.34 | 0.039 |
| Frequency | 1.24 | 0.99, 1.56 | 0.060 | 1.15 | 0.90, 1.46 | 0.270 |
| Voiding postponement | 0.94 | 0.85, 1.05 | 0.293 | 0.89 | 0.79, 1.00 | 0.055 |
| Voiding volume | 1.07 | 0.89, 1.29 | 0.489 | 1.00 | 0.82, 1.22 | 0.983 |
| **Exposure: logCRP (mg/L)** |  |  |  |  |  |  |
| UI (any) | 1.04 | 0.93, 1.17 | 0.449 | 1.03 | 0.91, 1.17 | 0.694 |
| Daytime wetting | 1.11 | 0.96, 1.30 | 0.166 | 1.06 | 0.89, 1.27 | 0.516 |
| Bedwetting | 0.99 | 0.85, 1.16 | 0.927 | 1.02 | 0.86, 1.21 | 0.839 |
| Urgency | 1.02 | 0.89, 1.15 | 0.814 | 0.96 | 0.84, 1.11 | 0.590 |
| Nocturia | 1.10 | 1.01, 1.21 | 0.034 | 1.06 | 0.95, 1.17 | 0.292 |
| Frequency | 1.12 | 0.96, 1.30 | 0.153 | 1.10 | 0.93, 1.30 | 0.273 |
| Voiding postponement | 0.99 | 0.91, 1.07 | 0.743 | 0.96 | 0.88, 1.04 | 0.323 |
| Voiding volume | 1.03 | 0.90, 1.17 | 0.694 | 1.01 | 0.87, 1.17 | 0.930 |

### Mediation results

Table 3 shows the TCE, NIE, NDE, and PM for model 1 (mediation through IL-6). There was weak evidence that the association between ACE score and any UI was mediated by IL-6 (OR_NIE_ 1·03, 95% CI 1·00-1·06, PM 21%). There was also weak evidence for a mediating effect of IL-6 between ACE score and daytime wetting (OR_NIE_ 1·06, 95% CI 1·01-1·10, PM 45%), bedwetting (OR_NIE_ 1·04, 95% CI 1·00-1·08, PM 31%), urgency (OR_NIE_ 1·03, 95% CI 1·00-1·07, PM 19%), and frequency (OR_NIE_ 1·04, 95% CI 1·00-1·08, PM 18%). There was no evidence of mediating effects of IL-6 on the association between ACE score and nocturia, voiding postponement, and voiding volume.

**Table 3.** Model 1 results for the mediation of adverse childhood experiences score and lower urinary tract symptoms via IL-6 in the imputed data sample (N = 4,745) OR odds ratio (per one unit increase in ACE score); CI confidence interval; UI urinary incontinence; NA not applicable; PM proportion mediated (%). Mediation models were fitted with IL-6 as the mediator of interest; age at blood draw, child BMI, emotional and behavioural problems, and constipation were considered intermediate confounders. All paths were adjusted for baseline confounders: sex, birthweight, gestational age, ethnicity, child developmental delay, maternal age, smoking during pregnancy, parity, maternal education, marital status, house tenure, and crowding. Analysis performed in 80 imputed datasets.

| ACE score | Total causal effect |  | Natural indirect effect |  | Natural direct effect |  | PM |
| --- | --- | --- | --- | --- | --- | --- | --- |
| Outcome | OR | 95% CI | OR | 95% CI | OR | 95% CI |  |
| UI (any) | 1.17 | 1.04, 1.31 | 1.03 | 1.00, 1.06 | 1.13 | 1.01, 1.28 | 21 |
| Daytime wetting | 1.14 | 0.98, 1.33 | 1.06 | 1.01, 1.10 | 1.08 | 0.92, 1.27 | 45 |
| Bedwetting | 1.15 | 0.98, 1.35 | 1.04 | 1.00, 1.08 | 1.10 | 0.93, 1.31 | 31 |
| Urgency | 1.19 | 1.06, 1.33 | 1.03 | 1.00, 1.07 | 1.15 | 1.02, 1.30 | 19 |
| Nocturia | 1.15 | 1.05, 1.25 | 1.01 | 0.99, 1.04 | 1.13 | 1.03, 1.24 | 8 |
| Frequency | 1.27 | 1.11, 1.45 | 1.04 | 1.00, 1.08 | 1.22 | 1.06, 1.40 | 18 |
| Voiding postponement | 1.09 | 1.01, 1.18 | 0.99 | 0.97, 1.02 | 1.10 | 1.01, 1.19 | NA |
| Voiding volume | 1.14 | 1.00, 1.31 | 1.03 | 0.99, 1.07 | 1.11 | 0.97, 1.28 | 23 |

There was no evidence of a mediating effect of CRP (model 2) on the associations between ACE score and LUTS (see Supplementary Table 11) after accounting for IL-6 as an intermediate confounder.

The results for the complete case analysis of the mediating effects of IL-6 (model 1) and CRP (model 2) on the association between ACE score and LUTS are in Supplementary Tables 12 and 13, respectively. The directions of effect and overall findings were similar.

## Discussion

Our study is the first to assess prospective associations between ACEs and LUTS in adolescence. An increased burden of ACEs (from birth to 8 years) was associated with an increase in the odds of LUTS in adolescence (at age 14). Our findings are consistent with earlier research that has reported an enduring effect of the total burden of ACEs on risk of LUTS in adults.^6,10,11^ Another novel aspect of our study is the examination of the prospective relationship between biomarkers of inflammation in childhood and LUTS in adolescence. We found that higher levels of the inflammatory biomarker IL-6 (but not CRP) at age 9 years were associated with increase odds of subsequent LUTS including any UI, daytime wetting and nocturia. This is also the first study to examine the mediating effect of inflammation as a possible biological mechanism that could underlie the relationship between ACEs and subsequent LUTS. We found weak evidence of a mediating effect of IL-6 (but not CRP) on the relationship between ACEs and LUTS.

### Strengths and limitations

Strengths of our study include the use of data from a large population-based birth cohort, the prospective design, availability of data on a range of self-reported LUTS in adolescence, and adjustment for a comprehensive set of confounders. ACEs were prospectively assessed at multiple timepoints from birth to age 8 years by multiple informants (mothers, partners, and children), compared with previous research which has relied on retrospective recall of ACEs. We assessed the total burden of ACEs, which is important because it is difficult to isolate the effect of a specific adverse event due to interrelationships between ACEs.^9^ We used adolescents’ self-reports of LUTS and did not restrict our analysis to those who met clinical diagnostic criteria. The study, therefore, provides evidence that ACEs are prospectively associated with LUTS in a non-clinical sample of adolescents in the community.

There are several limitations that should be considered when interpreting the current findings. Nonresponse and loss to follow-up occur more frequently among individuals who are exposed to socioeconomic disadvantage and ACEs.^18^ The complete-case sample had a lower proportion of participants with socioeconomic disadvantage and ACEs than the wider ALSPAC cohort, which may limit the internal validity of our findings. Restricting the analysis to the sample who provided complete data could cause bias, in addition to loss of statistical power due to a smaller sample size. We therefore used multiple imputation to address possible bias due to attrition and missing data. We reported the results from the analysis of the imputed data as the main findings and compared these with the results from the complete case analysis as recommended by Sterne et al.^19^

The ALSPAC cohort is predominantly white and affluent^14,15^ and hence we are unable to generalise our results to minority ethnic groups and less affluent populations. Further research in these underserved populations is vital to prevent widening inequalities in health research. Research in non-UK samples is also needed to examine if these findings generalise to children from other countries.

Inflammatory biomarkers were measured in non-fasting bloods at a single timepoint therefore, we do not have robust data on consistency of systemic levels of IL-6 and CRP within individuals over time. This is a potential limitation given the effects of age and puberty status on inflammation levels during childhood and adolescence.^20^ It is also possible that non-fasting blood samples of inflammatory biomarkers exhibit diurnal variation, resulting in that increased measurement error.^21^

Due to multiple testing, there is a possibility of an increase in type 1 errors. We do not, however, rely on p value thresholds to determine if results are statistically significant. Consistent with the recommended approach, we present effect estimates with their 95% confidence intervals in addition to the p-values.^22^

### Potential mechanisms that could explain the findings

ACEs have previously been described as getting *“under the skin”* and inducing *“physiological changes*”^23^ which can lead to an increased risk of chronic health problems. The physiological changes include an increased risk of systemic inflammation,^24^ which in turn, has been implicated as a risk factor for LUTS.^13^ There is evidence from animal models that exposure to chronic psychological stress and early life adversity can impact on lower urinary tract function due to functional and structural changes in the bladder, especially in the detrusor muscle and urothelium, and can lead to hypersensitivity of the afferent nerves in the bladder.^25^

We cannot rule out reverse causality as a possible explanation for the relationship between ACEs and LUTS. Persistent incontinence in children places a considerable burden on families and affects the quality of life of parents, and it is not uncommon for parents to adopt a negative coping style.^26^ Therefore, the child’s incontinence could be a cause, not a consequence, of adverse experiences.

Studies that examine inflammatory biomarker levels often exclude children with an infection that coincides with the time of blood collection because levels of IL-6 and CRP increase in response to acute infection but return to baseline afterwards. However, current infections could include UTIs; a known comorbid condition of LUTS.^27^ Exclusion of these individuals could, therefore, result in collider bias because LUTS are outcomes in the models.^28^ Consequently, we did not exclude individuals that reported any recent infection at blood sampling (N=446; 9·4%) nor those with a CRP level >10mg/L (N=57; 1·2%). A limitation of this is that some children will have temporarily inflated inflammation levels owing to other infection sources such as colds.

We found evidence of an association between IL-6 and LUTS, and a mediating effect of IL-6 on the association between ACE score and LUTS but found no evidence of effects with CRP. This is contrary to our hypothesis given that IL-6 upregulates the synthesis of CRP and both have functional roles in immunity.^29^ Despite their interrelationship IL-6 and CRP have distinct signalling pathways as well as functional roles in absence of inflammation. IL-6 is a pleiotropic cytokine operating through two distinct signalling pathways; classical and trans, the former of which is anti-inflammatory and results in the upregulation of CRP synthesis via signalling in hepatocytes.^30^ The trans-signalling pathway is pro-inflammatory, acts upon all cells, and is associated with chronic inflammatory conditions.^30^ It is plausible that mechanistically our results reflect the effects of IL-6 trans-signalling on risk of developing LUTS, independent of the classic signalling pathway which upregulates CRP levels.

## Conclusions

This study reports novel findings that could shed light on biological mechanisms that explain why ACEs increase the risk of subsequent LUTS. Further research is needed in other samples and with a wider range of inflammatory biomarkers to confirm our findings. Evidence of biological mechanisms linking ACEs to LUTS could lead to the identification of novel translational targets for intervention and potential therapeutic advances in the treatment of LUTS. ACEs can be difficult to modify, therefore, targeting the biological mechanisms could lead to secondary preventions aimed at reducing the risk of chronic LUTS.

Our findings should raise awareness amongst clinicians of the importance of screening for ACEs in children presenting with LUTS since they could be contributing factors. Screening for ACEs is also important because they can negatively affect treatment outcomes amongst children with LUTS.

## Data Availability

ALSPAC data are available by request from the ALSPAC Executive Committee for researchers who meet the criteria for access to confidential data (https://bristol.ac.uk/alspac/researchers/access/).

## Acknowledgements

We are extremely grateful to all the families who took part in this study, the midwives for their help in recruiting them, and the whole ALSPAC team, which includes interviewers, computer and laboratory technicians, clerical workers, research scientists, volunteers, managers, receptionists, and nurses.

This work is supported by funding from the Medical Research Council (grant ref: MR/V033581/1: Mental Health and Incontinence). GH, AGS, and JH are members of the MRC Integrative Epidemiology Unit at the University of Bristol (MC_UU_00011/7). AGS’ salary is funded by the European Union’s Horizon 2020 research and innovation programme under grant agreement No. 874739 (LongITools). The UK Medical Research Council and Wellcome (grant ref: 217065/Z/19/Z) and the University of Bristol provide core support for ALSPAC. This publication is the work of the authors and Kimberley Burrows will serve as guarantors for the contents of this paper. A comprehensive list of grants funding is available on the ALSPAC website: http://www.bristol.ac.uk/alspac/external/documents/grant-acknowledgements.pdf.

## Data sharing

ALSPAC data are available by request from the ALSPAC Executive Committee for researchers who meet the criteria for access to confidential data (https://bristol.ac.uk/alspac/researchers/access/). Code to conduct these analyses can be found on https://github.com/burrowsk upon publication.

