## Supplementary for "Adverse childhood experiences and lower urinary tract symptoms in adolescence: the mediating effect of inflammation"

**Supplementary Text**

Methods: parametric g-formula

We used the Stata package *gformula* to conduct parametric g-computation to estimate the total causal effect (TCE), natural direct effect (NDE) and the natural indirect effect (NIE).[1] G-formula uses Monte Carlo simulations to simulate the outcome, mediators and intermediate confounders under hypothetical interventions or “counter to the fact” scenarios (i.e. contrary to the intervention individuals actually received). The TCE is the value the outcome (e.g. any UI) would take if all individuals had been exposed to a one-unit increase in ACE score versus everyone not being exposed to a one unit increase in ACE score. The NDE is the direct (unmediated) effect of the exposure (e.g. ACE score) on the outcome (e.g. any UI) when the mediator (e.g. IL-6) takes the value it would be in absence of the exposure (i.e. ACE score of zero). More specifically it is modelled as the direct effect of exposure X = x+1 (e.g. an ACE score of 1) versus exposure X = x (e.g. an ACE score of 0) on outcome Y (e.g. any UI) if mediator M (e.g. IL-6) were set to whatever value it would be for X = x. The NIE is the effect of the exposure (e.g. ACE score) on the outcome (e.g. any UI) that operates by changing the mediator (e.g. IL-6). More specifically, it is modelled as the effect on outcome Y (e.g. any UI) if the exposure X = x+1 and the mediator M (e.g. IL-6) were changed from the value it would take if X = x to the value it would take if X = x+1. The proportion of the TCE that is mediated is calculated as $[OR_{NDE} (OR_{NIE} - 1)] / [OR_{NDE} \times OR_{NIE} - 1] \times 100$.[2]

Specifically for this analysis we used a Monte Carlo sample size of 100,000 to minimise fluctuations in effect estimates. Standard errors were estimated using 500 bootstrap samples. We also specified the command option *linexp* to specify that the exposure is continuous and its effect is assumed to be linear. For the imputed data we conducted mediation analysis for each of the 80 imputation datasets and output the results to log files. We used R to extract the log ORs and bootstrapped SEs for each of the TCE, NDE, and NIE from each log file. The mean of the log OR was computed and the SEs were calculated using Rubin’s rules.[3] The 95% confidence intervals were calculated from the combined log OR and SEs. Log OR (and 95% CIs) were exponentiated to represent ORs.

Results: description of sample

Supplementary Table 5 shows the descriptive statistics of study variables for the imputed and complete case samples. Any UI was reported by 5.3% of adolescents, with 4.6% and 4.5% of adolescents experiencing daytime wetting and bedwetting respectively. The most common LUTS were voiding postponement (15.6%) and nocturia (10.5%), while high frequency of urination was the least common (4.3%). All LUTS were moderately correlated (Rho 0.14-1.00) (see Supplementary Table 6). Prevalence of ACEs within the complete case data and imputed sample are given in Supplementary Table 7. In total 61.3% of individuals experienced at least one adversity between birth and 8 years of age within the imputed sample, whereas the prevalence in the complete case sample was 53.4%. Within the imputed sample 4.2% of individuals experienced ≥5 adversities. Due to sparsity of data for scores of seven or more, these scores were collapsed to represent a score of 7+. The most common adversity experienced was poor mental health or attempted suicide of parent(s) (36.2%), while the prevalence of sexual abuse was lowest at 1% within the imputed sample.

**Supplementary Figure 1 Directed acyclic graph (DAG) of mediation model 1 and model 2**

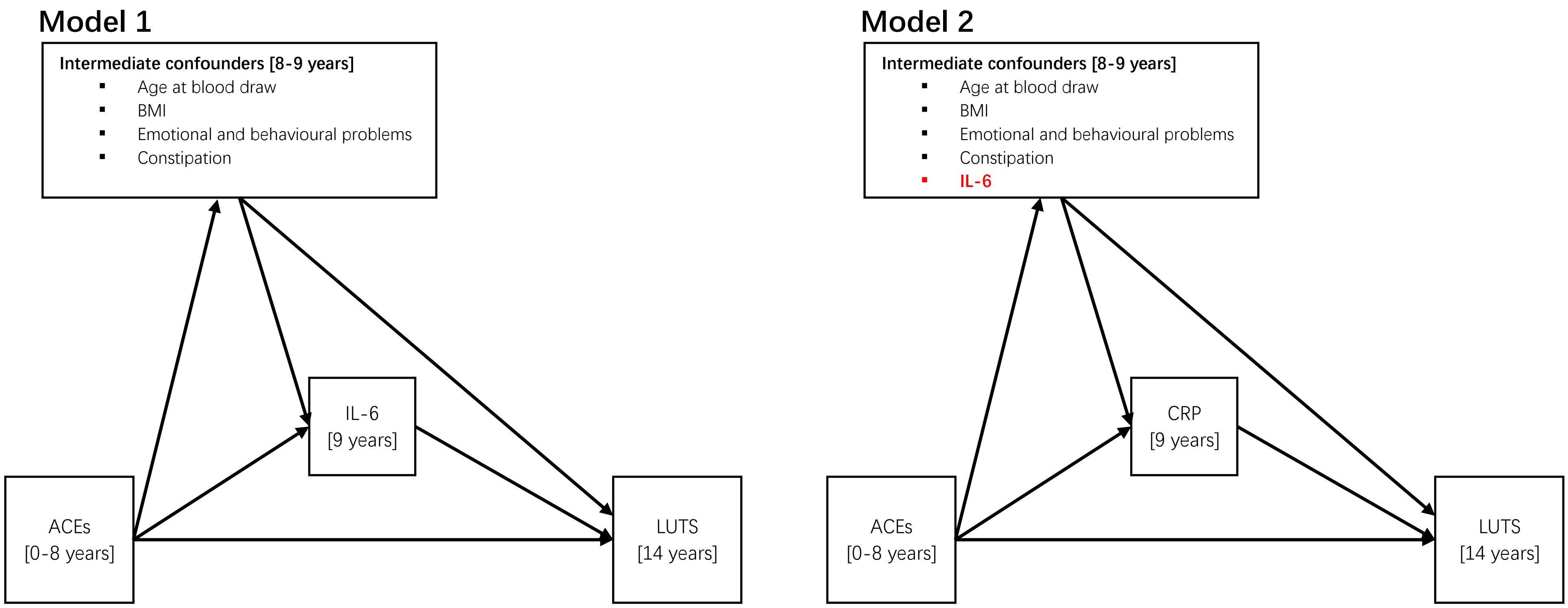

Mediation model 1 illustrates the mediating effect through IL-6; mediation model 2 illustrates the mediating effect through CRP and including IL-6 as an additional intermediate confounder. Each intermediate confounding path (age at blood draw, BMI, emotional and behavioural problems, and constipation (and Il-6 for model 2) were specified separately but are grouped together for clarity. For simplicity baseline confounders are omitted from the DAG as all confounders are proposed to cause all other model variables. ACEs adverse childhood experiences; LUTS lower urinary tract symptoms; BMI body mass index; CRP C-reactive protein; IL-6 interleukin 6.

**Supplementary Table 1 Variables and coding used to derive LUTS at age 14 years**

| Variable name | ALSPAC variable[1] | Question[2] | Responses | Final variable coding |
|---|---|---|---|---|
| ***How often does the following happen to you?*** | | | | |
| Daytime wetting | ccp550 | Wet yourself during the day? | 1: Never<br>2: Less than once per week<br>3: Once a week<br>4: 2-5 times a week<br>5: Nearly everyday<br>6: More than once a day<br>-10; -1: Missing | Recode:<br>1=0: No<br>2/6=1: Yes |
| Bed wetting | ccp551 | Wet the bed at night? | | |
| ***Over the last two weeks, how often have you...*** | | | | |
| Frequent urination | ccp521 | Had to go to the toilet for a wee more than 7 times a day? | 1: Never<br>2: A few times<br>3: Quite often<br>4: A lot<br>-10; -1: Missing | Recode:<br>1/2=0: Low frequency<br>3/4=1: High frequency |
| Nocturia | ccp527 | Woken up to go for a wee? | | |
| Urgency | ccp520 | Had a sudden feeling you need a wee and had to dash to the toilet? | | |
| Voiding postponement | ccp524 | Avoided going for a wee until the last moment because you were concentrating on other activities? | | |
| Low voided volume | ccp522 | Passed only a small amount when you went for a wee? | | |
| ***Urinary incontinence*** | | | | |
| Urinary incontinence UI | (derived) | Derived from ccp550 = Yes and/or ccp551 = Yes | 0: No<br>1: Yes | 0: No<br>1: Yes |

[1] ALSPAC variable names; these can be searched for using the ALSPAC variable search tool [http://variables.alspac.bris.ac.uk/]. [2] Data was collected via the ALSPAC questionnaire "Travelling, leisure and school"; For daytime and bedwetting, the young person was asked "Many of us have accidents sometimes. How often do the following happen to you?" responses included never, occasionally but less than once a week, about once a week, 2-5 times a week, nearly everyday, and more than once a day. For the other LUTS, the young person was asked "Over the last two weeks, how often have you…" responses included never, a few times, quite often, and a lot.

**Supplementary Table 2 Description of ACEs**

| ACE | Description | Number and age range[1] of questions asked | Respondents |
|---|---|---|---|
| **Sexual abuse** | Ever sexually abused, forced to perform sexual acts or touch someone in a sexual way | 7 [18 months – 7 years] | Mother |
| **Physical abuse** | Adult in family was ever physically cruel towards or hurt the child | 28 [0 – 7 years] | Mother and Partner |
| **Emotional abuse** | Parent was ever emotionally cruel towards the child or often said hurtful/insulting things to the child | 29 [0 – 7 years] | Mother and Partner |
| **Emotional neglect** | Child always felt excluded, misunderstood or never important to family, parents never asked or never listened when child talked about their free time | 1 [8 years] | Child completed |
| **Substance use** | Parent was a daily cannabis or any hard drug user, or, had an alcohol problem | 53 [8 weeks – 8 years] | Mother and Partner |
| **Mental health problems or suicide** | Parent was ever diagnosed with schizophrenia or hospitalised for a psychiatric problem, or, during the first 18 years of the child's life, parent had an eating disorder (bulimia or anorexia), used medication for depression or anxiety, attempted suicide or scored above previously established cut-offs for depression (Edinburgh Postnatal Depression Scale (EPDS) >12) | 50 [8 weeks – 8 years] | Mother and Partner |
| **Violence between parents** | Parents were ever affected by physically cruel behaviour by partner, or, ever violent towards each other, including hitting, choking, strangling, beating, shoving | 42 [8 months – 8 years] | Mother and Partner |
| **Parental separation** | Parents separated or divorced | 27 [8 months – 7 years] | Mother and Partner |
| **Bullying** | Child was a victim of bullying on a weekly basis | 1 [8 years] | Child completed |
| **Parent convicted** | Parent was convicted of a crime | 17 [8 weeks – 7 years] | Mother and Partner |

[1]The age of the index child at completion of the relevant questionnaire completed by either the mother, partner or child.

**Supplementary Table 3 Description and coding of confounders**

| Variable name | ALSPAC variable[1] | Measure | Responses | Final variable coding |
|---|---|---|---|---|
| **Child's sex assigned at birth** | | | | |
| Sex | kz021 | Recorded at birth by the fieldworkers who visited the maternity units | 1: Male<br>2: Female<br>-2; -1: Missing | Recode:<br>1=0: Male<br>2=1: Female |
| **Mother's age at birth** | | | | |
| Maternal age | mz028b | Derived variable: Grouped age of mother at delivery | Continuous (range <16 to >43 years)<br>-11; -10; -4; -2: Missing | Continuous (years) |
| **Parity** | | | | |
| Parity | b032 | Derived variable: number of previous pregnancies resulting in either a livebirth or a stillbirth | Continuous (range 0-22)<br>-7; -2; -1: Missing | Recode:<br>0=0: Nulliparous<br>1=1: One<br>2=1: Two<br>3/22=1: Three + |
| **Maternal education** | | | | |
| Maternal education | c645a | Derived variable: mum's highest educational qualification | 1: CSE/none<br>2: Vocational<br>3: O level<br>4: A level<br>5: Degree<br>-1: Missing | Recode:<br>4/5=0: A level or higher<br>3=1: O level<br>1/2=2: Vocational or less |
| **Home ownership** | | | | |
| Home ownership | a006 | Is your home:<br>-being bought/mortgaged<br>-owned - with no mortgage to pay<br>-rented from council<br>-rented from private landlord - furnished | 0: Mortgaged<br>1: Owned<br>2: Council rented<br>3: Rent private furnished<br>4: Rent private unfurnished | Recode:<br>0, 1, 3, 4=0: Owned/privately rented<br>2, 5, 6=1: HA/other |

| Variable name | ALSPAC variable[1] | Measure | Responses | Final variable coding |
|---|---|---|---|---|
| | | -rented from private landlord - unfurnished<br>-rented from housing association<br>-other (please describe) | 5: HA rented<br>6: Other | |
| **Child developmental level** | | | | |
| Adapted Denver Developmental Screening Test | kd685 | Derived variable: Total ALSPAC development score (prorated) | Continuous Z-score (range -1.4 to 3.3)<br>-102; -101: Missing | Continuous (Z-score) |
| **Ethnicity** | | | | |
| Child ethnic background | c804 | Derived variable: Ethnic group reported for the mother (respondent) or partner: Non-white if c801 or c802 had codes in the range of 2-9 | 1: White<br>2: Non-white<br>-1: Missing | Recode:<br>0: White<br>1: Non-white |
| **Birthweight** | | | | |
| Birthweight | kz030 | Derived variable: Preferred birthweight from multi-source:<br>Obstetric data, ALSPAC measurers, or birth notification | Continuous (range 200 to 5640 g)<br>-11; -10; -7; -2; -1: Missing | Continuous (g) |
| **Gestational age at birth** | | | | |
| Gestational age | bestgest | Derived variable: Best guess of gestation when pregnancy ended | Continuous (range 4 to 47 weeks)<br>-11; -10; -3; -2: Missing | Continuous (weeks) |
| **Marital status** | | | | |
| Marital status | a525 | What is your present marital status?<br>- Never married<br>- Widowed<br>- Divorced<br>- Separated<br>- Married (once only)<br>- Married for second or third time | 1: Never married<br>2: Widowed<br>3: Divorced<br>4: Separated<br>5: 1st marriage<br>6: Marriage 2 or 3<br>-1: Missing | Recode:<br>5/6=0: Married<br>1=1: Single<br>2/4=2: Divorced/separated/widowed |

| Variable name | ALSPAC variable[1] | Measure | Responses | Final variable coding |
|---|---|---|---|---|
| **Household crowding index** | | | | |
| Crowding index | a551 | Derived variable: Number of people in household (A550) divided by number of rooms (A045) | 1: <= 0.5<br>2: >0.5 - 0.75<br>3: >0.75 - 1<br>4: >1<br>-7; -1: Missing | Recode:<br>1/3=0: No crowding<br>4=1: Crowded |
| **Maternal smoking during pregnancy** | | | | |
| | b665 (1st trimester) | Did you smoke regularly in the first three months of pregnancy? | 1: No<br>2: Yes, cigarettes<br>3: Yes, cigars<br>5: Yes, other | Recode:<br>No (0) smoking during trimesters 1, 2 and 3 = **0**: "None" |
| | b667 (2nd trimester) | Did you smoke regularly in the last 2 weeks? | 1: No<br>2: Yes, cigarettes<br>3: Yes, cigars<br>5: Yes, other | Smoked during 1st trimester (2, 3, 5) but not the 2nd (0) and 3rd (0) = **1**: "Yes, quit early" |
| | c483 (3rd trimester) | How many cigarettes per day are you yourself smoking at the moment? | 0: None<br>1: 1-9<br>2: 10-19<br>3: 20+<br>-7: Missing | Smoked during trimesters 1 (2, 3, 5), 2 (2, 3, 5), and 3 (1, 2, 3) *or* either 1 and 2 or 1 and 3, or 2 and 3 = **2**: "Yes, throughout" |
| ***Intermediate confounders*** | | | | |
| **Body Mass Index (BMI)** | | | | |
| Clinical measures of height and weight (measured at mean age 8.65 years) | f8lf021a | Derived from measure of height (m) and weight (kg) at clinic visit. Calculated at $kg/m^2$ | Continuous score<br>-103; -102; -101: Missing | Continuous score |
| **Emotional and behavioural difficulties** | | | | |

| Variable name | ALSPAC variable[1] | Measure | Responses | Final variable coding |
|---|---|---|---|---|
| Adapted from Strengths and Difficulties Questionnaire (Eur Child Adolesc Psychiatry. 1998;7(3):125–30) (measured at mean age 8.26 years) | n8365f | Derived variable: SDQ total difficulties score (prorated) | Continuous score (range 0-40) -10; -6; -5: Missing | Continuous score |
| **Constipation** | | | | |
| Constipation (measured at mean age 8.73 years) | ks1037 | Child had constipation in past year | 1: Yes, saw doctor<br>2: Yes, did not see doctor<br>3: No, did not have<br>-1: Missing | Recode:<br>3=0: No<br>1/2=1: Yes |
| **Age of child at blood draw** | | | | |
| Age at blood draw (measured at mean age 9.88 years) | f9003c | Age of child's attendance to clinic | Continuous (range 105 to 140 months) | Continuous (months) |

[1] ALSPAC variable names; these can be searched for using the ALSPAC variable search tool [http://variables.alspac.bris.ac.uk/].

**Supplementary Table 4 Auxiliary variables for multiple imputation of missing data**

| | |
|---|---|
| Mother's opinion of neighbourhood during pregnancy | Household social class at 18 weeks gestation |
| Maternal depression score (EPDS) at 18 wks gestation | Daytime wetting between age 7 and 9 years |
| Mother divorced since pregnancy | Bedwetting between age 7 and 9 years |
| Mother became homeless during pregnancy | Frequent urinary between age 7 and 9 years |
| Difficulty affording food during pregnancy | Nocturia between age 7 and 9 years |
| Difficulty affording heating during pregnancy | Urgency between age 7 and 9 years |
| Maternal depression score (EPDS) at 32 wks gestation | Voiding postponement between age 7 and 9 years |
| Mother-reported highest educational level partner | Signs of urination between age 7 and 9 years |
| Maternal pre-pregnancy weight (Kg) | BMI at age 7 years |
| Maternal pre-pregnancy BMI | BMI at age 9 years |
| Antidepressant use by mother when child was 18yrs | Emotional and behavioural difficulties at age 6 years |
| Partner hard drug use during pregnancy | Constipation at age 7 years |
| Partner separated since pregnancy | |
| Partner convicted of an offence during pregnancy | |
| Partner depression score (EPDS) at 18 wks gestation | |
| Self-reported highest educational level partner | |
| Partner-reported highest educational level mother | |
| Maternal depression score (EPDS) when child was 18yrs | |
| Mother separated from partner when child was 18yrs | |
| Mother s partner was emotionally cruel when child was 18yrs | |
| Antidepressant use by mother when child was 18yrs | |
| Partner of child used physical force when child was 18-21yrs | |
| Partner of child used more severe physical force when child was 18-21yrs | |
| Partner of child have pressured them into kissing/touching when child was 18-21yrs | |
| Partner of child physically forced them into kissing/touching when child was 18-21yrs | |
| Partner of child used pressured them into sexual intercourse when child was 18-21yrs | |
| Partner of child physically forced them into sexual intercourse when child was 18-21yrs | |
| Partner of child made them feel scared of frightened when child was 18-21yrs | |

**Supplementary Table 5 Descriptive statistics of study variables for the imputed sample (N = 4,745) and complete case sample (N = 1,399)**

| Variable | Imputed sample (N = 4,745) | | Missing from eligible sample (N = 4,745)[2] | Complete case sample (N = 1,399) | |
|---|---|---|---|---|---|
| | Mean | SE | N [%] | Mean | SE |
| ACE score (0-10) | 1.30 | 0.02 | 1,996 [41.4] | 0.95 | 0.03 |
| IL-6 (pg/ml) | 1.28 | 0.02 | NA | 1.24 | 0.04 |
| CRP (mg/L) | 0.80 | 0.04 | NA | 0.71 | 0.05 |
| BMI at age 8 (kg/m$^2$) | 17.06 | 0.03 | 955 [20.1] | 16.93 | 0.06 |
| [1]SDQ total difficulties score (0-40) | 7.70 | 0.08 | 867 [18.3] | 6.91 | 0.13 |
| Age at blood draw (years) | 9.87 | 0.01 | NA | 9.78 | 0.01 |
| Child development level (Z-score) | 0.22 | 0.02 | 1,226 [25.8] | -0.01 | 0.02 |
| Birthweight (kg) | 3.44 | 0.01 | 56 [1.2] | 3.44 | 0.01 |
| Gestational age at delivery (weeks) | 39.51 | 0.03 | NA | 39.53 | 0.05 |
| Parity (n) | 0.81 | 0.01 | 163 [3.4] | 0.74 | 0.02 |
| Maternal age (years) | 29.12 | 0.07 | NA | 29.77 | 0.11 |
| | % | SE | N [%] | % | SE |
| Childs sex (female) | 49.29 | 0.73 | NA | 50.68 | 1.34 |
| Ethnicity (non-white) | 4.48 | 0.32 | 216 [4.6] | 2.14 | 0.39 |
| Urinary incontinence (any) | 8.24 | 0.54 | 1,276 [26.9] | 5.29 | 0.60 |
| Daytime wetting | 4.59 | 0.44 | 1,277 [26.9] | 3.57 | 0.50 |
| Bedwetting | 4.49 | 0.42 | 1,276 [26.9] | 2.57 | 0.42 |
| Urgency | 6.40 | 0.47 | 1,271 [26.8] | 5.00 | 0.58 |
| Frequency (high) | 4.34 | 0.46 | 1,281 [27.0] | 2.22 | 0.39 |
| Nocturia | 10.51 | 0.55 | 1,288 [27.1] | 7.58 | 0.71 |
| Voiding volume (low) | 5.68 | 0.46 | 1,292 [27.2] | 3.79 | 0.51 |
| Voiding Postponement | 15.61 | 0.63 | 1,280 [27.0] | 12.58 | 0.89 |
| Constipation | 10.26 | 0.50 | 729 [15.4] | 9.15 | 0.77 |
| Maternal smoking during pregnancy | | | | | |
| None | 80.56 | 0.58 | 89 [1.9] | 86.63 | 0.91 |
| Yes, quit early | 5.11 | 0.33 | | 4.00 | 0.52 |
| Yes, throughout | 14.33 | 0.51 | | 9.36 | 0.78 |
| Maternal education | | | | | |
| A-level or higher | 43.63 | 0.73 | 159 [3.4] | 52.54 | 1.34 |
| O level | 34.77 | 0.70 | | 34.38 | 1.27 |
| Vocational or less | 21.61 | 0.61 | | 13.08 | 0.90 |
| Martial status | | | | | |
| Married | 81.72 | 0.57 | 101 [2.1] | 87.85 | 0.87 |
| Single | 13.74 | 0.51 | | 8.22 | 0.73 |
| Divorced/separated/widowed | 4.54 | 0.31 | | 3.93 | 0.52 |
| Housing tenure (rented or other) | 12.06 | 0.48 | 143 [3.0] | 6.15 | 0.64 |
| Crowding (>1) | 4.48 | 0.32 | 204 [4.3] | 2.29 | 0.40 |

ACE adverse childhood experience; BMI body mass index; SDQ strengths and difficulties questionnaire; CRP C-reactive protein; IL6 Interleukin-6. 80 imputed datasets. The complete case sample comprises individuals with data for ACEs, inflammation, LUTS, all confounders and intermediate confounders. [1] Emotional and behavioural problems. [2] total number of missing participants for each variable. NA none missing

**Supplementary Table 6 Tetrachoric correlations between urinary incontinence/LUTS (eligible sample data N = 3,377)**

| | Urinary incontinence (any) | Daytime wetting | Bedwetting | Urgency | Nocturia | Frequency | Voiding postponement | Voiding volume |
|---|---|---|---|---|---|---|---|---|
| Urinary incontinence (any) | | | | | | | | |
| Daytime wetting | **1.00** | | | | | | | |
| Bedwetting | **1.00** | **0.57** | | | | | | |
| Urgency | **0.31** | **0.37** | **0.23** | | | | | |
| Nocturia | **0.24** | **0.27** | **0.29** | **0.40** | | | | |
| Frequency | **0.29** | **0.32** | **0.28** | **0.54** | **0.45** | | | |
| Voiding postponement | **0.30** | **0.35** | **0.22** | **0.37** | **0.30** | **0.34** | | |
| Voiding volume | **0.19** | 0.14 | **0.17** | **0.42** | **0.31** | **0.40** | **0.43** | |

Bolded values are correlations with P value <0.05

**Supplementary Table 7 Distributions of adverse childhood experiences within imputed data sample (N = 4,745) and complete case sample (N = 1,399)**

| Adverse childhood experiences (ACE) | Observed – eligible sample (N = 4,745) | Observed – complete case sample (N = 1,399) | Imputed[1] (N= 4,745) |
|---|---|---|---|
| | % [SE] | % [SE] | % [SE] |
| **ACE Score** | | | |
| 0 | 45.30 [0.94] | 46.60 [1.33] | 38.71 [0.79] |
| 1 | 28.43 [0.86] | 28.38 [1.21] | 27.00 [0.72] |
| 2 | 14.00 [0.66] | 14.01 [0.93] | 15.29 [0.63] |
| 3 | 7.59 [0.50] | 6.65 [0.67] | 9.50 [0.52] |
| 4 | 3.13 [0.33] | 3.29 [0.48] | 5.26 [0.40] |
| 5 | 1.33 [0.22] | 0.93 [0.26] | 2.79 [0.32] |
| 6 | 0.18 [0.08] | <0.36 [NA][2] | 1.00 [0.18] |
| 7+ | <0.18 [NA][2] | <0.36 [NA][2] | 0.49 [0.15] |
| *Missing[3] (N = 1,966; 41.4 %)* | | | |
| **Individual ACEs (% missing[3])** | | | |
| Sexual abuse (7.1) | 0.43 [0.10] | 0.36 [0.16] | 1.00 [0.20] |
| Physical abuse (17.5) | 6.23 [0.39] | 7.00 [0.68] | 7.93 [0.48] |
| Emotional abuse (26.5) | 15.49 [0.61] | 13.94 [0.94] | 18.71 [0.73] |
| Emotional neglect (20.9) | 1.81 [0.22] | 1.57 [0.33] | 4.19 [0.44] |
| Substance abuse (18.2) | 8.40 [0.45] | 7.80 [0.72] | 10.54 [0.54] |
| Poor mental health or suicide (17.4) | 33.47 [0.75] | 30.38 [1.23] | 36.17 [0.77] |
| Violence between parents (27.6) | 17.61 [0.65] | 15.94 [0.98] | 22.04 [0.72] |
| Parental separation (18.0) | 16.38 [0.59] | 11.29 [0.85] | 19.43 [0.67] |
| Bullying (20.8) | 1.54 [0.20] | 1.64 [0.34] | 3.74 [0.44] |
| Convicted of offences (16.3) | 5.26 [0.35] | 5.15 [0.59] | 6.64 [0.45] |

[1] 80 datasets imputed; [2] There are five or fewer participants within the ACE score count; [3] % with missing data for individual ACEs within the eligible sample; the complete case sample comprises individuals with data for ACEs, inflammation, LUTS, all confounders and intermediate confounders.

**Supplementary Table 8 Association results between adverse childhood experiences score and inflammation in the imputed sample (N = 4,745)**

| | Univariable | | | Multivariable | | |
|---|---|---|---|---|---|---|
| **Outcome** | **exp(Beta)** | **95% CI** | **p-value** | **exp(Beta)** | **95% CI** | **p-value** |
| ***Exposure: ACE Score*** | | | | | | |
| logIL-6 | 1.04 | 1.02, 1.06 | <0.001 | 1.03 | 1.01, 1.05 | 0.002 |
| logCRP | 1.02 | 0.99, 1.05 | 0.18 | 1.01 | 0.98, 1.04 | 0.47 |
| ***Exposure: logIL-6*** | | | | | | |
| logCRP | 1.97 | 1.90, 2.04 | <0.001 | 1.78 | 1.72, 1.84 | <0.001 |

CI confidence interval; CRP C-reactive protein; IL6 Interleukin-6. Multivariable models adjusted for sex, birthweight, gestational age, ethnicity, child developmental delay, maternal age, smoking during pregnancy, parity, maternal education, marital status, house tenure, and crowding. The associations between IL-6 and CRP are additionally adjusted for age at blood draw, child BMI, emotional and behavioural problems, and constipation and ACE score. exp(Beta) represents the percentage change in the outcome per unit increase in exposure. Log is the natural logarithm.

**Supplementary Table 9 Associations between adverse childhood experiences score and lower urinary tract symptoms in the complete case data sample (N = 1,399)**

| ACE score | Univariable | | | Multivariable | | |
|---|---|---|---|---|---|---|
| **Outcome** | **OR** | **95% CI** | **p-value** | **OR** | **95% CI** | **p-value** |
| UI (any) | 1.14 | 0.94, 1.36 | 0.163 | 1.13 | 0.93, 1.36 | 0.205 |
| Daytime wetting | 1.13 | 0.89, 1.39 | 0.297 | 1.07 | 0.83, 1.35 | 0.562 |
| Bedwetting | 1.24 | 0.95, 1.56 | 0.090 | 1.30 | 0.99, 1.67 | 0.046 |
| Urgency | 1.21 | 1.00, 1.44 | 0.042 | 1.21 | 1.00, 1.46 | 0.048 |
| Nocturia | 1.08 | 0.92, 1.27 | 0.331 | 1.06 | 0.89, 1.24 | 0.530 |
| Frequency | 1.08 | 0.80, 1.42 | 0.583 | 1.21 | 0.88, 1.61 | 0.213 |
| Voiding postponement | 1.04 | 0.90, 1.18 | 0.595 | 1.02 | 0.89, 1.17 | 0.734 |
| Voiding volume | 1.19 | 0.96, 1.45 | 0.104 | 1.15 | 0.91, 1.42 | 0.217 |

OR odds ratio; CI confidence interval; UI urinary incontinence; multivariable models adjusted for baseline confounders (sex, birthweight, gestational age, ethnicity, child developmental delay, maternal age, smoking during pregnancy, parity, maternal education, marital status, house tenure, and crowding).

**Supplementary Table 10 Associations between inflammatory biomarkers and lower urinary tract symptoms in the complete case sample (N = 1,399)**

| Inflammation | **Univariable** | | | **Multivariable** | | |
|---|---|---|---|---|---|---|
| **Outcome** | **OR** | **95% CI** | **p-value** | **OR** | **95% CI** | **p-value** |
| ***logIL-6 (pg/ml)*** | | | | | | |
| UI (any) | 1.34 | 1.04, 1.73 | 0.026 | 1.35 | 1.03, 1.77 | 0.027 |
| Daytime wetting | 1.65 | 1.21, 2.24 | 0.001 | 1.54 | 1.11, 2.12 | 0.010 |
| Bedwetting | 1.11 | 0.77, 1.59 | 0.575 | 1.20 | 0.83, 1.74 | 0.347 |
| Urgency | 1.07 | 0.82, 1.39 | 0.636 | 0.99 | 0.75, 1.31 | 0.931 |
| Nocturia | 1.27 | 1.02, 1.58 | 0.030 | 1.22 | 0.97, 1.53 | 0.086 |
| Frequency | 1.12 | 0.76, 1.66 | 0.555 | 1.16 | 0.78, 1.70 | 0.465 |
| Voiding postponement | 0.92 | 0.77, 1.09 | 0.324 | 0.87 | 0.72, 1.05 | 0.138 |
| Voiding volume | 1.10 | 0.82, 1.49 | 0.533 | 1.03 | 0.75, 1.43 | 0.834 |
| ***logCRP (mg/L)*** | | | | | | |
| UI (any) | 1.09 | 0.90, 1.30 | 0.385 | 1.10 | 0.89, 1.34 | 0.364 |
| Daytime wetting | 1.18 | 0.94, 1.46 | 0.132 | 1.09 | 0.85, 1.38 | 0.489 |
| Bedwetting | 1.04 | 0.78, 1.35 | 0.771 | 1.12 | 0.83, 1.48 | 0.441 |
| Urgency | 1.02 | 0.83, 1.24 | 0.859 | 0.95 | 0.75, 1.19 | 0.679 |
| Nocturia | 1.10 | 0.93, 1.28 | 0.251 | 1.04 | 0.87, 1.23 | 0.673 |
| Frequency | 1.11 | 0.82, 1.45 | 0.483 | 1.14 | 0.83, 1.52 | 0.384 |
| Voiding postponement | 0.90 | 0.78, 1.03 | 0.138 | 0.85 | 0.73, 1.00 | 0.049 |
| Voiding volume | 1.10 | 0.88, 1.36 | 0.371 | 1.10 | 0.85, 1.40 | 0.453 |

OR odds ratio; CI confidence interval; CRP C-reactive protein; IL-6 Interleukin-6; UI urinary incontinence; multivariable models adjusted for baseline confounders (sex, birthweight, gestational age, ethnicity, child developmental delay, maternal age, smoking during pregnancy, parity, maternal education, marital status, house tenure, and crowding), intermediate confounders (age at blood draw, child BMI, emotional and behavioural problems, and constipation), and ACE score. Log is the natural logarithm.

**Supplementary Table 11 Model 2 results for the mediation of adverse childhood experiences score and lower urinary tract symptoms via CRP in the imputed data sample (N = 4,745)**

| ACE score | **Total causal effect** | | **Natural indirect effect** | | **Natural direct effect** | | |
|---|---|---|---|---|---|---|---|
| **Outcome** | **OR** | **95% CI** | **OR** | **95% CI** | **OR** | **95% CI** | **PM** |
| UI (any) | 1.18 | 1.05, 1.33 | 1.00 | 0.97, 1.03 | 1.18 | 1.05, 1.33 | 0.00 |
| Daytime wetting | 1.18 | 1.01, 1.38 | 0.99 | 0.95, 1.03 | 1.19 | 1.03, 1.39 | NA |
| Bedwetting | 1.19 | 1.01, 1.40 | 0.98 | 0.94, 1.02 | 1.22 | 1.04, 1.42 | NA |
| Urgency | 1.20 | 1.07, 1.35 | 1.00 | 0.96, 1.03 | 1.20 | 1.07, 1.36 | NA |
| Nocturia | 1.16 | 1.06, 1.27 | 1.00 | 0.97, 1.02 | 1.16 | 1.06, 1.27 | NA |
| Frequency | 1.32 | 1.16, 1.51 | 0.99 | 0.95, 1.04 | 1.33 | 1.17, 1.52 | NA |
| Voiding postponement | 1.11 | 1.02, 1.20 | 1.00 | 0.98, 1.03 | 1.10 | 1.02, 1.19 | 4.39 |
| Voiding volume | 1.17 | 1.03, 1.34 | 1.00 | 0.96, 1.04 | 1.17 | 1.03, 1.34 | 0.00 |

OR odds ratio; CI confidence interval; UI urinary incontinence; NA not applicable; PM proportion mediated (%). Mediation models were fitted with CRP as the mediator of interest; age at blood draw, child BMI, emotional and behavioural problems, and constipation, and IL-6 were considered intermediate confounders. All paths were adjusted for baseline confounders: sex, birthweight, gestational age, ethnicity, child developmental delay, maternal age, smoking during pregnancy, parity, maternal education, marital status, house tenure, and crowding. Analysis performed in 80 imputed datasets.

**Supplementary Table 12 Results for the mediation of adverse childhood experiences score and lower urinary tract symptoms via IL-6 in the complete case sample (N = 1,399)**

| ACE score | **Total causal effect** | | **Natural indirect effect** | | **Natural direct effect** | | |
|---|---|---|---|---|---|---|---|
| **Outcome** | **OR** | **95% CI** | **OR** | **95% CI** | **OR** | **95% CI** | PM |
| UI (any) | 1.13 | 0.95, 1.35 | 1.05 | 1.06, 1.09 | 1.07 | 0.85, 1.25 | 43 |
| Daytime wetting | 1.05 | 0.92, 1.31 | 1.04 | 1.04, 1.08 | 1.01 | 0.80, 1.18 | 80 |
| Bedwetting | 1.07 | 0.99, 1.30 | 1.03 | 1.03, 1.04 | 1.14 | 1.05, 1.44 | 20 |
| Urgency | 1.23 | 1.10, 1.59 | 1.03 | 1.02, 1.04 | 1.19 | 1.06, 1.50 | 16 |
| Nocturia | 1.07 | 0.91, 1.28 | 1.03 | 1.02, 1.05 | 1.03 | 0.86, 1.20 | 51 |
| Frequency | 1.03 | 0.96, 1.12 | 1.01 | 1.01, 1.02 | 1.02 | 0.95, 1.08 | 34 |
| Voiding postponement | 1.03 | 0.88, 1.18 | 1.00 | 0.98, 1.04 | 1.02 | 0.87, 1.18 | 0.00 |
| Voiding volume | 1.20 | 1.10, 1.52 | 1.02 | 1.01, 1.06 | 1.17 | 1.07, 1.52 | 12 |

OR odds ratio; CI confidence interval (bias-corrected and accelerated 95% CI's); UI urinary incontinence; PM proportion mediated (%). Mediation models were fitted with IL-6 as the mediator of interest; age at blood draw, child BMI, emotional and behavioural problems, and constipation were intermediate confounders. All paths were adjusted for baseline confounders: sex, birthweight, gestational age, ethnicity, child developmental delay, maternal age, smoking during pregnancy, parity, maternal education, marital status, house tenure, and crowding.

**Supplementary Table 13 Model 2 results for the mediation of adverse childhood experiences score and lower urinary tract symptoms via CRP in the complete case sample (N = 1,399)**

| ACE score | **Total causal effect** | | **Natural indirect effect** | | **Natural direct effect** | | |
|---|---|---|---|---|---|---|---|
| **Outcome** | **OR** | **95% CI** | **OR** | **95% CI** | **OR** | **95% CI** | PM |
| UI (any) | 1.16 | 1.01, 1.46 | 0.98 | 0.95, 1.00 | 1.18 | 1.05, 1.44 | NA |
| Daytime wetting | 1.07 | 0.95, 1.41 | 0.97 | 0.95, 0.98 | 1.10 | 1.00, 1.51 | NA |
| Bedwetting | 1.18 | 1.10, 1.40 | 0.99 | 0.97, 1.00 | 1.19 | 1.12, 1.39 | NA |
| Urgency | 1.24 | 1.12, 1.57 | 0.99 | 0.95, 1.01 | 1.25 | 1.13, 1.55 | NA |
| Nocturia | 1.06 | 0.90, 1.04 | 0.99 | 0.94, 1.01 | 1.07 | 0.93, 1.30 | NA |
| Frequency | 1.04 | 0.98, 1.17 | 0.99 | 0.99, 1.00 | 1.05 | 0.99, 1.23 | NA |
| Voiding postponement | 1.16 | 1.03, 1.56 | 0.98 | 0.95, 0.99 | 1.19 | 1.07, 1.53 | NA |
| Voiding volume | 1.04 | 0.93, 1.20 | 1.00 | 0.97, 1.02 | 1.07 | 0.93, 1.21 | NA |

OR odds ratio; CI confidence interval (bias-corrected and accelerated 95% CI's); UI urinary incontinence; NA not applicable; PM proportion mediated (%). Mediation models were fitted with CRP as the mediator of interest; age at blood draw, child BMI, emotional and behavioural problems, and constipation, and IL-6 were intermediate confounders. All paths were adjusted for baseline confounders: sex, birthweight, gestational age, ethnicity, child developmental delay, maternal age, smoking during pregnancy, parity, maternal education, marital status, house tenure, and crowding.

**STROBE Statement—checklist of items that should be included in reports of observational studies**

| | Item No | Recommendation |
|---|---|---|
| **Title and abstract**<br>**See abstract** | 1 | (*a*) Indicate the study's design with a commonly used term in the title or the abstract |
| | | (*b*) Provide in the abstract an informative and balanced summary of what was done and what was found |
| **Introduction** | | |
| Background/rationale<br>**Introduction paragraphs 1 & 2** | 2 | Explain the scientific background and rationale for the investigation being reported |
| Objectives<br>**Introduction paragraph 3, Figure 2, Supplementary Figure 1** | 3 | State specific objectives, including any prespecified hypotheses |
| **Methods** | | |
| Study design<br>**Methods** | 4 | Present key elements of study design early in the paper |
| Setting<br>**Methods: sample** | 5 | Describe the setting, locations, and relevant dates, including periods of recruitment, exposure, follow-up, and data collection |
| Participants<br>**Methods: sample, Figure 1** | 6 | (*a*) *Cohort study*—Give the eligibility criteria, and the sources and methods of selection of participants. Describe methods of follow-up<br>*Case-control study*—Give the eligibility criteria, and the sources and methods of case ascertainment and control selection. Give the rationale for the choice of cases and controls<br>*Cross-sectional study*—Give the eligibility criteria, and the sources and methods of selection of participants |
| | | (*b*) *Cohort study*—For matched studies, give matching criteria and number of exposed and unexposed<br>*Case-control study*—For matched studies, give matching criteria and the number of controls per case |
| Variables<br>**Methods, Supplementary Tables 1-4** | 7 | Clearly define all outcomes, exposures, predictors, potential confounders, and effect modifiers. Give diagnostic criteria, if applicable |
| Data sources/ measurement<br>**Supplementary Tables 1-4** | 8* | For each variable of interest, give sources of data and details of methods of assessment (measurement). Describe comparability of assessment methods if there is more than one group |
| Bias<br>**Methods: Statistical analysis and missing data** | 9 | Describe any efforts to address potential sources of bias |
| Study size<br>**Methods: sample, Figure 1** | 10 | Explain how the study size was arrived at |
| Quantitative variables<br>**Methods: statistical analysis** | 11 | Explain how quantitative variables were handled in the analyses. If applicable, describe which groupings were chosen and why |
| Statistical methods<br>**Methods: statistical analysis, Supplementary Text** | 12 | (*a*) Describe all statistical methods, including those used to control for confounding |
| | | (*b*) Describe any methods used to examine subgroups and interactions |
| | | (*c*) Explain how missing data were addressed |
| | | (*d*) *Cohort study*—If applicable, explain how loss to follow-up was addressed<br>*Case-control study*—If applicable, explain how matching of cases and controls was addressed<br>*Cross-sectional study*—If applicable, describe analytical methods taking account of sampling strategy |
| | | (*e*) Describe any sensitivity analyses |

| | | |
|---|---|---|
| **Results** | | |
| Participants<br>**Figure 1, Supplementary Text** | 13* | (a) Report numbers of individuals at each stage of study—eg numbers potentially eligible, examined for eligibility, confirmed eligible, included in the study, completing follow-up, and analysed |
| | | (b) Give reasons for non-participation at each stage |
| | | (c) Consider use of a flow diagram |
| Descriptive data<br>**Supplementary Text, Supplementary Tables 5-7** | 14* | (a) Give characteristics of study participants (eg demographic, clinical, social) and information on exposures and potential confounders |
| | | (b) Indicate number of participants with missing data for each variable of interest |
| | | (c) *Cohort study*—Summarise follow-up time (eg, average and total amount) |
| Outcome data<br>**Supplementary Text, Supplementary Tables 5-7** | 15* | *Cohort study*—Report numbers of outcome events or summary measures over time |
| | | *Case-control study*—Report numbers in each exposure category, or summary measures of exposure |
| | | *Cross-sectional study*—Report numbers of outcome events or summary measures |
| Main results<br>**Results, Tables 1-3, Supplementary Tables 8-13** | 16 | (*a*) Give unadjusted estimates and, if applicable, confounder-adjusted estimates and their precision (eg, 95% confidence interval). Make clear which confounders were adjusted for and why they were included |
| | | (*b*) Report category boundaries when continuous variables were categorized |
| | | (*c*) If relevant, consider translating estimates of relative risk into absolute risk for a meaningful time period |
| Other analyses<br>**NA** | 17 | Report other analyses done—eg analyses of subgroups and interactions, and sensitivity analyses |
| **Discussion** | | |
| Key results<br>**Discussion paragraph 1** | 18 | Summarise key results with reference to study objectives |
| Limitations<br>**Discussion: Strengths and limitations** | 19 | Discuss limitations of the study, taking into account sources of potential bias or imprecision. Discuss both direction and magnitude of any potential bias |
| Interpretation<br>**Discussion: potential mechanisms** | 20 | Give a cautious overall interpretation of results considering objectives, limitations, multiplicity of analyses, results from similar studies, and other relevant evidence |
| Generalisability<br>**Discussion: Strengths and limitations** | 21 | Discuss the generalisability (external validity) of the study results |
| **Other information** | | |
| Funding<br>**Funding section** | 22 | Give the source of funding and the role of the funders for the present study and, if applicable, for the original study on which the present article is based |

*Give information separately for cases and controls in case-control studies and, if applicable, for exposed and unexposed groups in cohort and cross-sectional studies.

**Note:** An Explanation and Elaboration article discusses each checklist item and gives methodological background and published examples of transparent reporting. The STROBE checklist is best used in conjunction with this article (freely available on the Web sites of PLoS Medicine at http://www.plosmedicine.org/, Annals of Internal Medicine at http://www.annals.org/, and Epidemiology at http://www.epidem.com/). Information on the STROBE Initiative is available at www.strobe-statement.org.
